# The effect of International Monetary Fund programmes on population health

**DOI:** 10.1101/2021.03.12.21253458

**Authors:** Elias Nosrati, Michael Marmot, Lawrence P. King

## Abstract

**Background:** As one of the world’s most powerful international financial organisations, the International Monetary Fund (IMF) is uniquely positioned to shape global developmental trajectories by influencing domestic policy arrangements. However, its role in shaping population health is understudied.

**Methods:** We use previously unavailable cross-national time-series data and an instrumental-variables method to examine the causal impact of the IMF’s structural adjustment programmes on age-standardised all-cause mortality rates and disability-adjusted life years (DALYs) in 187 countries between 1990 and 2017. For this purpose, we specify two-way fixed-effects regression models.

**Findings:** According to our baseline models, IMF programmes cause 392 excess all-cause deaths (95% CI: 176–608; *p* = 0.0004) and 4,205 excess DALYs (95% CI: 2,429–5,981; *p* = 0.000004) per 100,000 population. This aggregate effect appears to be driven by IMF-mandated privatisation reforms, which lead to 562 excess all-cause deaths (95% CI: 267–857; *p* = 0.0002) and 5,285 excess DALYs (95% CI: 2,749–7,822; *p* = 0.00004) per 100,000 population.

**Interpretation:** Structural adjustment programmes play a significant role in perpetuating preventable disability and death in developing contexts. The IMF’s policy reforms, especially those pertaining to the privatisation of state-owned enterprises, increase excess mortality rates and disease burdens in the world’s most vulnerable regions.

**Funding:** None.

## Introduction

In its capacity as global lender of last resort, as provider of technical assistance, and as agent of economic surveillance, the International Monetary Fund (IMF) is uniquely positioned to shape the developmental trajectories of countries experiencing financial distress through its conditional lending programmes.^1,2^ In particular, its power to influence its clients’ domestic policy arrangements leads the IMF to play a pivotal role in moulding the institutional determinants of population health, especially through the articulation of social and economic policies.^3^ However, despite the politically controversial nature of the Fund’s global interventions, robust empirical evidence surrounding the causal relation between so-called “structural adjustment programmes” and population health remains scarce. The purpose of this article is to address this gap by using previously unavailable data and a compound instrumental variables technique to derive unbiased causal effect estimates.

In times of financial turbulence, the IMF extends credit to debt-ridden yet fiscally constrained nations in exchange for wide-ranging policy reforms — known as “conditionalities” — that are intended to ensure macroeconomic stability and good governance. Previous studies have posited that such conditionalities, by prioritising debt repayment over long-term public investments and by promoting liberal economic policies, have the potential to influence population health outcomes, for instance by reducing state capacity and eroding healthcare systems via fiscal austerity and the privatisation of state-owned enterprises.^4–7^ Three mechanisms have been identified as central to this process.^3^ First, the IMF-mandated pursuit of macroeconomic targets via extensive spending cuts and institutional reforms can directly affect healthcare infrastructures, including the quality of provision, the extent of coverage, and the day-to-day functioning of individual care units.^8,9^ Second, health systems can be indirectly affected by interventions in non-health-related policy domains, such as currency devaluation, that increases the price of imported medicines and equipment.^10^ Third, structural adjustment programmes — by redefining a wide gamut of institutional parameters and pathways — can act upon and activate the social determinants of health, including (un)employment, poverty, and reduced education expenditures.^3,6,11^

Previous research has demonstrated strong associations between IMF programmes and a variety of health outcomes, yet causal evidence remains scarce.^6^ We seek to offer the first systematic analysis of how structural adjustment affects age-standardised all-cause mortality rates and disability-adjusted life years across 187 countries between 1990 and 2017. Using a compound instrumentation method suited to isolating exogenous treatment variation, we find statistically robust and substantively large effects of IMF intervention on our outcomes of interest. Moreover, by drawing on earlier scholarship linking rapid privatisation policies to ill health, we demonstrate that much of this aggregate effect appears to be driven by privatisation conditionalities. These have the potential to increase disease and death burdens by fuelling unemployment and work-related insecurity, by curtailing the public provision of various social benefits — such as housing, healthcare, and social services — and by causing broader economic disruption and reduced state administrative capacity.^12,13^

**RESEARCH IN CONTEXT**

#### Evidence before this study

The International Monetary Fund’s (IMF) structural adjustment programmes have been the subject of extensive public attention and debate, but little is known about how these programmes affect population health in low- and middle-income countries. We searched PubMed, Science Direct, Scopus, Web of Science, and Google Scholar from database inception until 20 September 2020, using various combinations of the terms “International Monetary Fund”, “IMF”, “structural adjustment”, “health”, “mortality”, and “disease”. We found a number of articles discussing the linkages between IMF programmes and health outcomes, yet only a handful of studies provided plausible causal evidence. Most previous analyses are also limited in their temporal scope, geographical focus, or chosen health outcome. To our knowledge, there are no original empirical investigations of the causal relation between structural adjustment and age-standardised all-cause mortality rates or disability-adjusted life years (DALYs).

#### Added value of this study

We empirically assessed the impact of structural adjustment programmes on population health by using previously unavailable panel data from 187 countries between 1990 and 2017. To derive causal effect estimates, we drew on recent methodological advances in the evaluation of IMF programmes by constructing an instrumental variable both for programme participation and for policy-specific conditionalities. We discovered statistically robust and substantively large adverse effects on age-standardised all-cause mortality and DALY burdens per 100,000 population. Our study provides the first comprehensive overview of the consequences of IMF programmes for health and, more generally, it adds to the scientific understanding of the distal determinants of population health.

#### Implications of all the available evidence

IMF-mandated policy reforms, especially those geared towards rapid privatisation, are harmful to population health and contribute to the persistence of high mortality and disease burdens in developing contexts. The role of international multilateral organisations such as the IMF in shaping population health outcomes should be taken seriously by researchers and policy-makers.

## Data and methods

We use two alternative outcome variables: the annual age-standardised all-cause mortality rate and the burden of disability-adjusted life years (DALYs) per 100,000 population in 187 non-high income countries between 1990 and 2017. These variables are available from the Institute for Health Metrics and Evaluation.^14,15^ We employ two sets of treatment variables to assess the effects of structural adjustment, both of which are drawn from the newly released IMF Monitor Database.^16^ On the one hand, we use a dichotomous indicator of whether a country is under an IMF programme to estimate the average effect of IMF intervention. On the other hand, to further probe the specific nature of structural loan conditions and their relation to the outcome variables, we assess the role of IMF-mandated privatisations of state-owned enterprises, as motivated by earlier scholarship linking rapid privatisation reforms to substantially deteriorating health outcomes.^12,13^

Our control variables include a series of macroeconomic fundamentals that can influence the propensity of countries with varying health profiles to participate in IMF programmes. These are gross domestic product (GDP) per capita, measured in constant 2010 US dollars,^17^ a binary financial crisis indicator,^18^ and foreign reserves in months of imports.^17^ We also control for a number of political factors, including a general democracy index^19^ and a more refined measure of egalitarian democracy;^20^ a coup d’état indicator^21^ as a measure of political instability; and United Nations General Assembly (UNGA) voting alignment with the G7 countries.^22^ The latter variable is construed as a proxy for geo-strategic alignment and is known to be predictive of IMF programme participation and potentially of the types of conditionalities received by borrowing countries.^23^ Finally, we also control for average years of completed education in the female population aged 25–29.^24^ Descriptive statistics are provided in Table 1.

**Table 1:** Descriptive statistics

| STATISTIC | N | MEAN | ST. DEV. | MIN | MAX |
| --- | --- | --- | --- | --- | --- |
| All-cause mortality rate | 5,460 | 1,023 | 514 | 301 | 13,041 |
| All-cause disability-adjusted life years | 5,460 | 3,522 | 3,978 | 209 | 26,224 |
| IMF programme | 5,165 | 0.3 | 0.5 | 0.0 | 1.0 |
| IMF-mandated privatisation | 5,131 | 0.1 | 0.5 | 0.0 | 12.0 |
| GDP per capita | 4,733 | 11,561 | 17,103 | 164 | 111,968 |
| Financial crisis | 5,236 | 0.1 | 0.2 | 0.0 | 1.0 |
| Foreign reserves | 3,932 | 4.4 | 4.7 | 0.002 | 79 |
| Democracy | 4,909 | 3.5 | 6.6 | -10 | 10 |
| Egalitarian democracy | 4,097 | 0.4 | 0.2 | 0.03 | 0.9 |
| Coup d'état | 4,396 | 0.02 | 0.1 | 0.0 | 1.0 |
| UNGA voting alignment | 5,012 | -1.6 | 1.0 | -3.9 | 1.4 |
| Mean years of female education | 5,404 | 8.9 | 4.0 | 0.5 | 16 |

NOTES: The all-cause mortality rate is age-standardised. Both the all-cause mortality rate and the disability-adjusted life years are per 100,000 population. The second column lists the number of observed country-years. The privatisation variable counts the total number of privatisation conditionalities imposed on a borrowing country by the IMF. The general democracy index ranges from -10 to 10. The egalitarian democracy index ranges from 0 to 1.

To empirically examine the causal relation between structural adjustment and population health, we posit the following data-generating process:

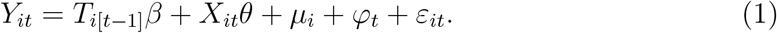

*Y*_*it*_ denotes one of the two alternative outcome variables as measured in country *i* at time *t*; *T*_*i*[*t−*1]_ is a dichotomous indicator of IMF programme participation, lagged by one year to allow for delayed effects to manifest; *X* is a vector of control variables; *µ* captures time-invariant country-specific effects; *φ* measures time-fixed effects; and *ε* is a stochastic error term. Our principal quantity of interest is *β*, which is a causal effect parameter to be estimated. However, in an observational study such as this, parameter estimates are likely to suffer from endogeneity bias. For example, perhaps countries that have poor political leadership are both more likely to make economic policy mistakes that result in an IMF loan programme, and are also more likely to exercise poor leadership on public health policy, resulting in excess mortality. This is visualised in Figure 1, where some unmeasured confounder *C* can render the estimated association between *T* and *Y* biased, even after adjusting for observed covariates *X*.

**Figure 1:**
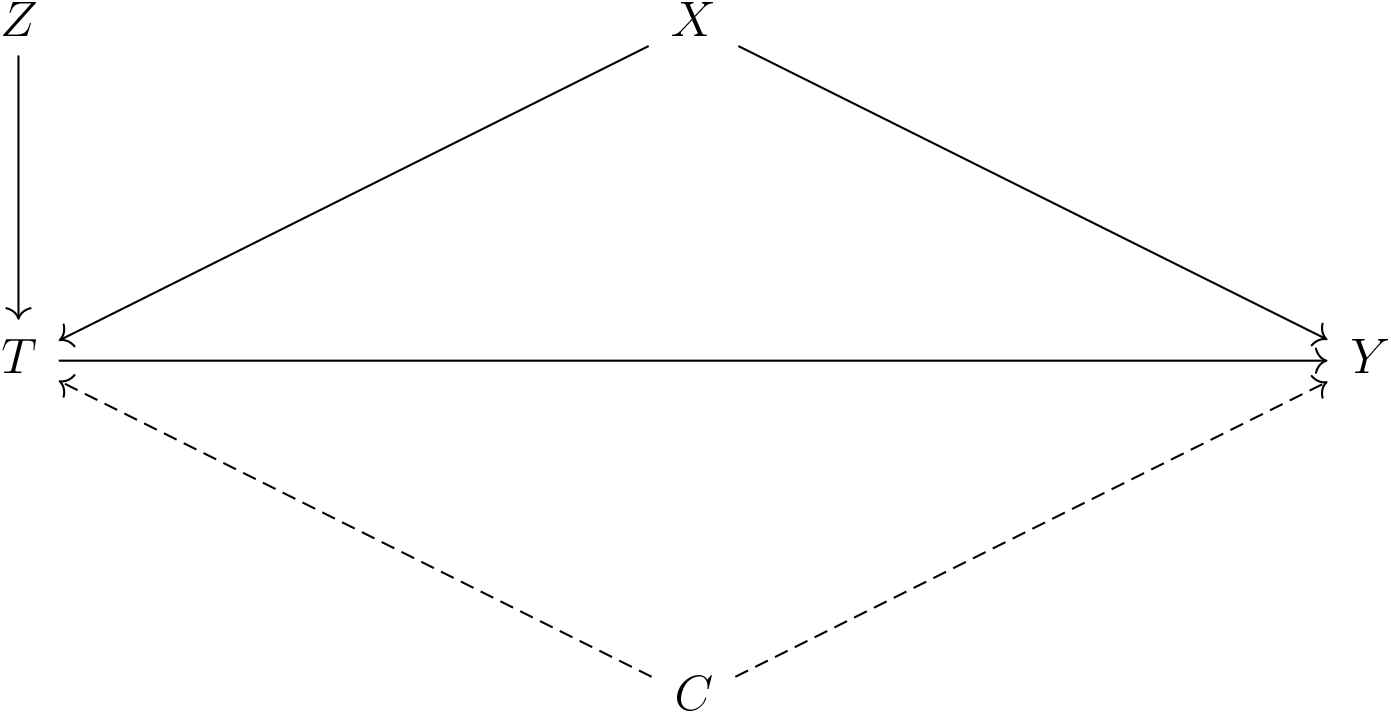
Causal graph depicting the effect of the treatment variable (*T* = IMF programme) on the outcome (*Y* = all-cause mortality rate or DALY burden), identified via a compound instrument (*Z* = country-specific average IMF programme participation *×* annual IMF liquidity constraint), net of both measured covariates (*X*) and unmeasured confounders (*C*).

One possible solution to this issue is to construct an instrumental variable, *Z*, which is correlated with the treatment but uncorrelated with any other variables in the causal system, thereby isolating exogenous variation in *T*. To do this, we follow recent methodological advances in the study of IMF programmes^25,26^ by adopting a compound instrument derived from the interaction between the country-specific average exposure to IMF programmes over the sample period and the number of countries with an IMF programme in a given year, which approximates the Fund’s annual budget constraint.^27^ We argue that the instrument is exogenous — i.e., capable of capturing causal effects — because unit-specific changes in structural adjustment that deviate from a country’s long-run average exposure to IMF programmes are generated only by IMF decisions that are unrelated to any given country’s health profile. We unpack and justify this argument as follows. A valid instrument is one that affects the causal variable of interest (IMF programmes) but one that is also uncorrelated with potential confounders. The IMF’s annual budget constraint should meet this criterion: it is clearly related to IMF programmes, but in general it should be uncorrelated with country-specific health profiles. In addition, previous research has shown that the IMF is more likely to impose harsher loan conditions when it faces liquidity concerns, regardless of the client country.^25^ However, some countries might be more likely to seek the IMF’s assistance than other countries, and this might generate a spurious association between structural adjustment programmes and population health outcomes. To address this concern, we add country- and time-fixed effects to our model, which helps net out any systematic propensity to enter IMF programmes, as well as any aggregate time trends (such as overall trends in life expectancy). Thus, the joint usage of an instrumental variable and of country- and time-fixed effects provides a rigorous framework for causal identification. We nevertheless provide a comprehensive sensitivity analysis that quantifies any residual confounding, as detailed below.

In short, the chosen instrument is relevant to the treatment insofar as liquidity concerns lead the IMF to impose more stringent loan conditions, yet it isolates exogenous variation in that annual budget constraints operate independently of any given country. Our identifying assumption is therefore that the outcome of interest in countries with different propensities to participate in IMF programmes will not be affected by changes in the Fund’s budgetary constraint other than through the impact of structural adjustment: that is, as per Figure 1, the only pathway linking *Z* to *Y* goes via *T*. We thus obtain a two-stage regression model with the following selection equation:

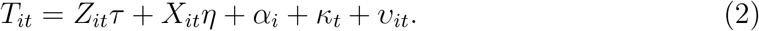

We then re-specify the model in equation (1) as follows, with 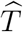being a vector of fitted values from equation (2):

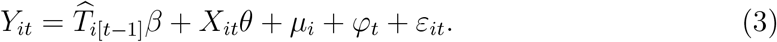

To empirically assess the strength of the chosen instrument, we compare the model in equation (2) to a restricted first-stage regression in which the effect *τ* of *Z* on *T* is set to be null, obtaining a *χ*^2^ test statistic of 53.641 on 1 degree of freedom (*p <* 0.001). Hence *Z* comfortably satisfies the benchmark for identifying a strong instrument. We control for the endogenous relation between *T* and *Y* potentially induced by any time-invariant propensity of countries with a prior health disadvantage to select into IMF programmes by adjusting for country-fixed effects, whereas year-fixed effects help account for broader aggregate changes that affect all countries simultaneously. All variance estimators are consistent with serial autocorrelation, heteroskedasticity, and country-level clustering effects. All analyses are conducted in R, version 4.0.2.

Given that we cannot empirically verify that our instrument is strictly exogenous, the persistence of unmeasured residual confounding is possible. To address this concern, we conduct a simple non-parametric sensitivity analysis that allows us to quantify the amount of unmeasured confounding that would in theory be required to eliminate our estimated causal effect 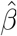. As per Figure 1, let *C* denote an unmeasured confounder. Then the bias factor, *ℬ*, is defined as the difference between 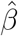 and what 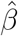 would have been had we controlled for *C* as well. Assuming *C* is binary, we define

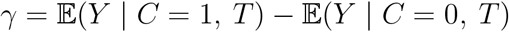

as the net effect of the unmeasured confounder on the outcome. We also define

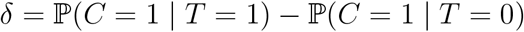

as the difference in the prevalence of the unmeasured confounder between the treatment and control groups. Then the bias factor is the product of these two sensitivity parameters: *ℬ* = *γ × δ*.^28,29^ In assessing the sensitivity of our model coefficients to unmeasured confounding, we ask how large *γ* would have to be in order to reduce our estimated effect size 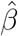 to zero. We address this question by visualising how *ℬ* changes as the two sensitivity parameters (co-)vary across a range of possible values.

## Results

Outputs from the two-way fixed-effects regression models are displayed in Tables 2 and 3 for the aggregate impact of IMF programmes on all-cause mortality and disability-adjusted life years. Our baseline models, shown in the first rows of each table, suggest that the adoption of IMF programmes in one year causes 392 excess all-cause deaths (95% CI: 176–608; *p* = 0.0004) and 4,205 excess DALYs (95% CI: 2,429–5,981; *p* = 0.000004) per 100,000 population in the following year. To assess the robustness of these parameter estimates to additional covariates, we introduce our control variables. However, to avoid multicollinearity issues and the loss of too many observations at once due to missing data, we add and remove these controls one by one and inspect the corresponding change in the treatment coefficient. We find that structural adjustment remains a robust predictor of variation in both health outcomes, but especially for disability-adjusted life years. For both outcome variables, the greatest attenuation in the estimated effect — by up to roughly one-fourth — occurs when controlling for foreign reserves, which is a strong predictor of selection into IMF programmes, and (egalitarian) democracy, which is associated with lower disease and death burdens. As we will discuss below, this attenuation may plausibly reflect a mediating pathway from structural adjustment to population health via political equality and participation.

**Table 2:** IMF programmes and all-cause mortality rates

| CONTROL VARIABLE | CONTROL COEFFICIENT | IMF<sub>*t*-1</sub> COEFFICIENT |
| --- | --- | --- |
| — | — | 392***<br>(110) |
| Log of GDP per capita | −85<br>(75) | 363**<br>(123) |
| Financial crisis | −41<br>(26) | 400***<br>(112) |
| Foreign reserves | 5**<br>(2) | 291**<br>(112) |
| Democracy | −7*<br>(3) | 355***<br>(106) |
| Egalitarian democracy | −280<br>(214) | 266*<br>(116) |
| Coup d'état | 156<br>(145) | 360*<br>(140) |
| UNGA voting alignment | −8<br>(18) | 371***<br>(110) |
| Female education | −13<br>(16) | 402***<br>(110) |

NOTES: The outcome variable is the annual age-standardised all-cause mortality rate per 100,000 population between 1990 and 2017. Each row is a separate two-way fixed-effects regression wherein the effect of IMF programmes on all-cause mortality is adjusted for the control variable listed in the first column. All models are also adjusted for country- and time-fixed effects. The IMF variable, lagged by one year, is instrumented as described in the DATA AND METHODS section. The corresponding parameter estimate is interpreted as the excess number of deaths per 100,000 population caused by structural adjustment. Standard errors consistent with serial autocorrelation, heteroskedasticity, and unit clustering are shown in parentheses below each parameter estimate. Statistical significance levels: \* $p < 0.05$ ; \*\* $p < 0.01$ ; \*\*\* $p < 0.001$ .

**Table 3:** IMF programmes and disability-adjusted life years

| CONTROL VARIABLE | CONTROL COEFFICIENT | IMF<sub>*t*-1</sub> COEFFICIENT |
| --- | --- | --- |
| — | — | 4,205***<br>(906) |
| Log of GDP per capita | -1,132<br>(771) | 3,632***<br>(1,009) |
| Financial crisis | -289<br>(247) | 4,265***<br>(920) |
| Foreign reserves | 0.15<br>(15) | 2,992***<br>(792) |
| Democracy | -78*<br>(32) | 3,761***<br>(841) |
| Egalitarian democracy | -6,280**<br>(1,978) | 3,918***<br>(1,009) |
| Coup d'état | 427<br>(290) | 4,216***<br>(1,224) |
| UNGA voting alignment | 67<br>(183) | 4,033***<br>(110) |
| Female education | -105<br>(175) | 4,297***<br>(878) |

NOTES: The outcome variable is the annual burden of disability-adjusted life years per 100,000 population between 1990 and 2017. Each row is a separate two-way fixed-effects regression wherein the effect of IMF programmes is adjusted for the control variable listed in the first column. All models are also adjusted for country- and time-fixed effects. The IMF variable, lagged by one year, is instrumented as described in the DATA AND METHODS section. The corresponding parameter estimate is interpreted as the excess number of disability-adjusted life years per 100,000 population caused by structural adjustment. Standard errors consistent with serial autocorrelation, heteroskedasticity, and unit clustering are shown in parentheses below each parameter estimate. Statistical significance levels: \* $p < 0.05$ ; \*\* $p < 0.01$ ; \*\*\* $p < 0.001$ .

Given that our control variables may be difficult to interpret in their own right — especially insofar as some of them may plausibly mediate or modify the causal relation between structural adjustment and population health — we proceed to the sensitivity analysis described above, the results for which are visualised in Figure 2. The X-axis ranges from 0 to 1, with higher values indicating a higher prevalence of the confounder in the treatment group (i.e., in countries with IMF programmes), whilst the Y-axis denotes the magnitude of the net effect of *C* on the *Y* that would be required to completely eliminate the effect of structural adjustment on the outcome. In light of the argument concerning the exogeneity of our chosen instrument, we believe it is plausible that the amount of residual confounding remains moderate. As such, the most likely values of *δ* would be at the lower end of the X-axis in Figure 2. At, say, *δ* = 0.1, *C* would have to cause nearly 4,000 excess deaths and over 40,000 excess DALYs per 100,000 population to nullify the effect of structural adjustment, which seems implausible. For the sake of argument, assume that the bias factor is roughly one-tenth of 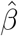 — i.e., *ℬ* = 39 in cause of all-cause mortality and *ℬ* = 420 in the case of DALYs. Then the bias-adjusted effect of IMF programmes would still be 353 excess all-cause deaths (95% CI: 137– 569; *p <* 0.001) and 3,785 excess DALYs (95% CI: 2,009–5,561; *p <* 0.001). Overall, the sensitivity analysis suggests that an inordinate amount of unmeasured confounding would be needed to cast doubt upon our causal estimates.

**Figure 2:**
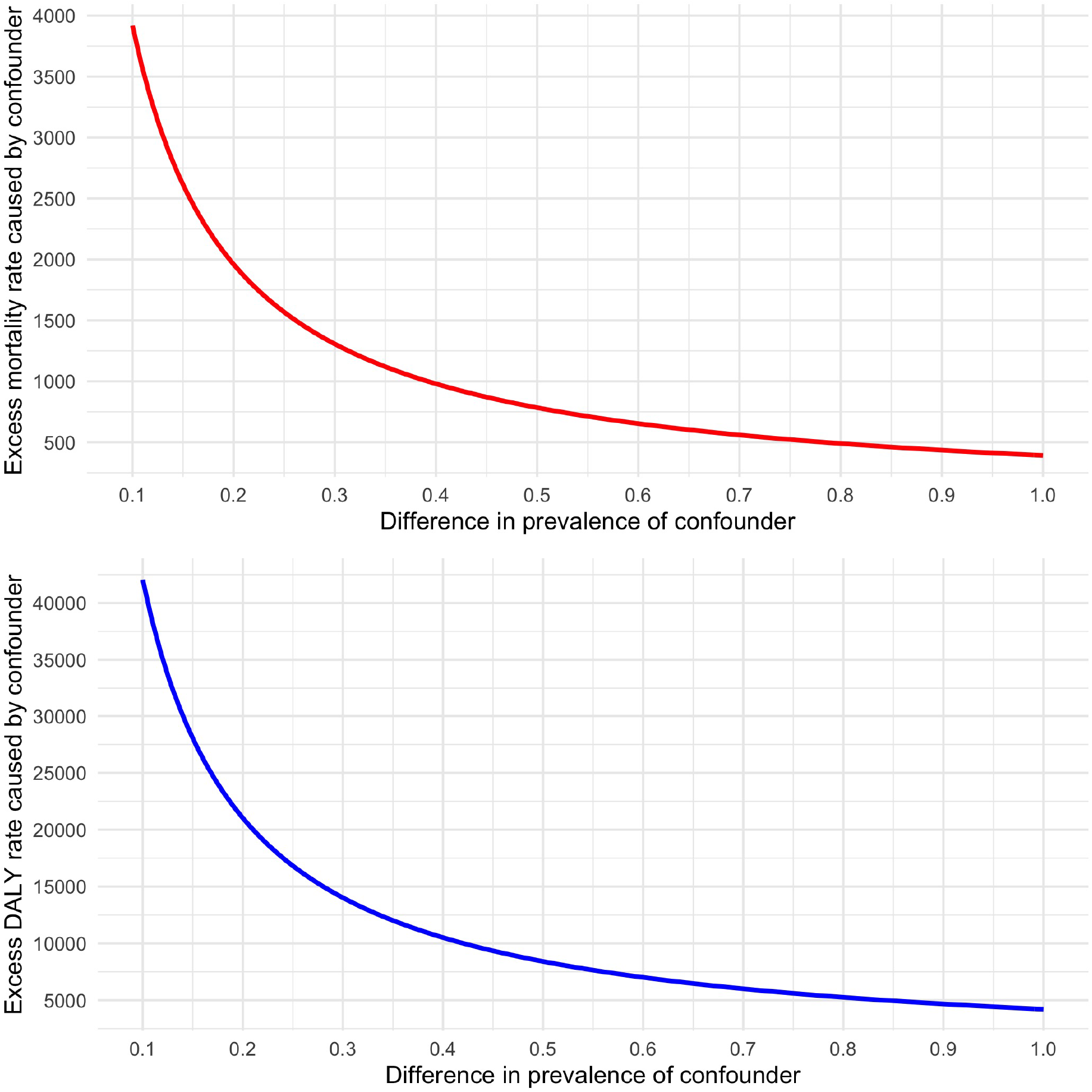
Sensitivity analysis plot to assess residual confounding of the estimated effect 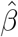 of IMF programmes on population health as per Tables 2 and 3. Values of *δ* (X-axis) and *γ* (Y-axis) that lie on the solid lines would completely eliminate the estimated effects of structural adjustment. Values above the plotted curves would reverse the sign of the estimated effects.

We proceed to the analysis of IMF-mandated privatisation reforms, for which we dichotomise the privatisation measure for ease of interpretation. We also verify that the chosen instrument — now defined as the interaction between the country-specific average exposure to privatisation policies and the Fund’s annual budget constraint — passes the required significance threshold, obtaining a *χ*^2^ test statistic of 40.464 on 1 degree of freedom (*p <* 0.001). The results of this analysis are displayed in Tables 4 and 5. According to the baseline model shown in the first row of each table, privatisation measures in one year lead to 562 excess all-cause deaths (95% CI: 267–857; *p* = 0.0002) and 5,285 excess DALYs (95% CI: 2,749–7,822; *p* = 0.00004) per 100,000 population in the following year. These estimates are robust to additional controls, though the effect sizes are attenuated by adjusting for egalitarian democracy and the incidence of coups d’état in the first model, and by foreign reserves in the second model. However, the substantive finding — that privatisation conditionalities significantly affect health outcomes — remains unaltered. Figure 3 shows the results of the corresponding sensitivity analysis, once again suggesting that very high levels of bias are required to eliminate our estimated causal effects. For instance, if *δ* = 0.2, *C* would have to cause close to 3,000 excess deaths and 30,000 excess DALYs in order to nullify 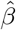.

**Table 4:** IMF-mandated privatisation reforms and all-cause mortality rates

| CONTROL VARIABLE | CONTROL COEFFICIENT | PRIVATISATION<sub>*t*-1</sub> COEFFICIENT |
| --- | --- | --- |
| — | — | 562***<br>(150) |
| Log of GDP per capita | -238**<br>(74) | 515**<br>(178) |
| Financial crisis | -6<br>(20) | 563***<br>(150) |
| Foreign reserves | 5**<br>(2) | 580**<br>(213) |
| Democracy | -3<br>(3) | 518***<br>(152) |
| Egalitarian democracy | -81<br>(204) | 430*<br>(168) |
| Coup d'état | 164<br>(151) | 427*<br>(172) |
| UNGA voting alignment | -1<br>(16) | 530***<br>(153) |
| Female education | -7<br>(15) | 572***<br>(152) |

NOTES: The outcome variable is the annual age-standardised all-cause mortality rate per 100,000 population between 1990 and 2017. Each row is a separate two-way fixed-effects regression wherein the effect of privatisation conditionalities is adjusted for the control variable listed in the first column. All models are also adjusted for country- and time-fixed effects. The conditionality variable, lagged by one year, is dichotomised and instrumented as described in the DATA AND METHODS section. The corresponding parameter estimate is interpreted as the excess number of all-cause deaths per 100,000 population caused by IMF-mandated privatisation reforms. Standard errors consistent with serial autocorrelation, heteroskedasticity, and unit clustering are shown in parentheses below each parameter estimate. Statistical significance levels: \* $p < 0.05$ ; \*\* $p < 0.01$ ; \*\*\* $p < 0.001$ .

**Table 5:** IMF-mandated privatisation reforms and disability-adjusted life years

| CONTROL VARIABLE | CONTROL COEFFICIENT | PRIVATISATION<sub>*t*-1</sub> COEFFICIENT |
| --- | --- | --- |
| — | — | 5,285***<br>(1,294) |
| Log of GDP per capita | -2,486***<br>(752) | 4,671**<br>(1,565) |
| Financial crisis | 131<br>(172) | 5,254***<br>(1,290) |
| Foreign reserves | -2<br>(17) | 2,992**<br>(1,718) |
| Democracy | -41<br>(28) | 4,803***<br>(1,320) |
| Egalitarian democracy | -3,754**<br>(1,428) | 4,596**<br>(1,556) |
| Coup d'état | 397<br>(237) | 4,536**<br>(1,626) |
| UNGA voting alignment | 73<br>(174) | 5,009***<br>(1,308) |
| Female education | -32<br>(175) | 5,332***<br>(1,294) |

NOTES: The outcome variable is the burden of disability-adjusted life years per 100,000 population between 1990 and 2017. Each row is a separate two-way fixed-effects regression wherein the effect of privatisation conditionalities is adjusted for the control variable listed in the first column. All models are also adjusted for country- and time-fixed effects. The conditionality variable, lagged by one year, is dichotomised and instrumented as described in the DATA AND METHODS section. The corresponding parameter estimate is interpreted as the excess number of disability-adjusted life years per 100,000 population caused by IMF-mandated privatisation reforms. Standard errors consistent with serial autocorrelation, heteroskedasticity, and unit clustering are shown in parentheses below each parameter estimate. Statistical significance levels: \* $p < 0.05$ ; \*\* $p < 0.01$ ; \*\*\* $p < 0.001$ .

**Figure 3:**
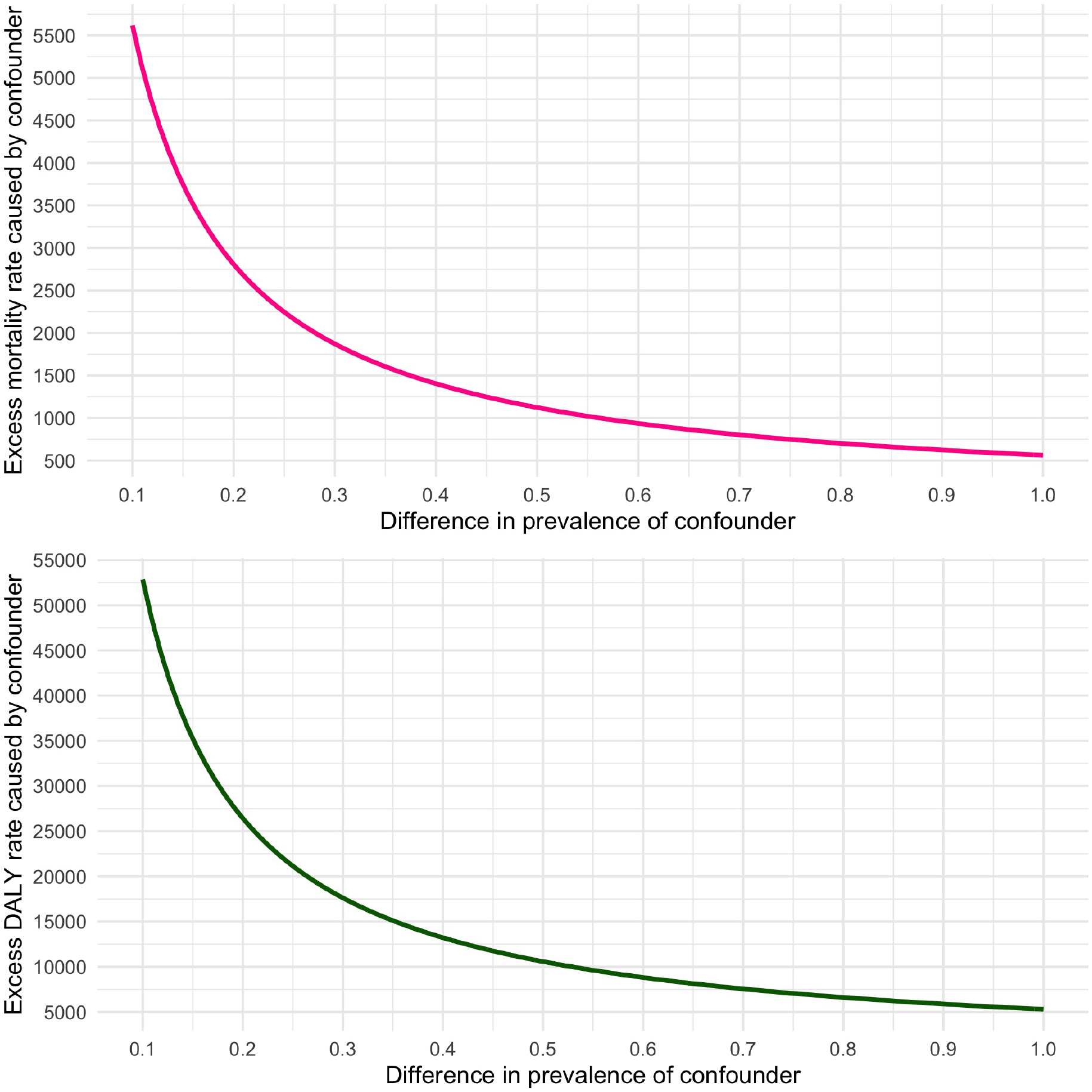
Sensitivity analysis plot to assess residual confounding of the estimated effect 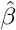 of IMF-mandated privatisation reforms on population health as per Tables 4 and 5. Values of *δ* (X-axis) and *γ* (Y-axis) that lie on the solid lines would completely eliminate the estimated effects of structural adjustment. Values above the plotted curves would reverse the sign of the estimated effects.

## Discussion

Our analysis provides novel causal evidence from previously unavailable cross-national time-series data, linking the IMF’s interventions in low- and middle-income countries to poor health outcomes. We corroborate earlier studies and hypotheses surrounding this topic, yet we offer new empirical insights. Although we are unable to probe the specific mechanisms by which the estimated causal effects take place, we rely on the extant literature that has highlighted three likely causal pathways from structural adjustment to population health. These involve pathways affecting the healthcare system both directly and indirectly, as well as pathways operating via the social determinants of health.^3^ In the latter case, the rapid privatisation of state-owned enterprises has previously been linked to higher unemployment and fractured labour markets, heightened social insecurity and stress, the concentrated loss of various social benefits, and the erosion of public institutions.^12,13^ Against this backdrop, we find that IMF programmes as a whole, and especially privatisation conditionalities, have strong adverse impacts on both age-standardised all-cause mortality rates and on the total burden of disability-adjusted life years.

We note that the estimated effect of structural adjustment is most strongly attenuated by controlling for foreign reserves, which is a central economic predictor of programme participation, but also by democratic politics. On the one hand, this may be viewed as evidence of estimation bias. On the other hand, in the latter case, it is also possible to consider a mediating pathway whereby structural adjustment shapes population health by eroding political equality and participation. Previous research has shown that IMF programmes impose concentrated losses on key social groups, undermine bureaucratic quality, and thereby incentivise corruption and deepen inequality.^30–32^ These factors shape the nature of democratic politics which, in turn, influences population health outcomes.^33–35^ As such, we surmise that the average causal effect of structural adjustment may plausibly operate via a range of mechanisms — including via a client country’s political infrastructure and the attendant quality of democratic institutions.

We acknowledge the limitations of our analysis. Our relatively limited sample size — though substantially larger than those of many earlier studies — may have reduced our statistical power to probe various quantities of interest and may have increased the estimation uncertainty accompanying our models. Nonetheless, our principal findings remain remarkably robust and are highly significant, both statistically and substantively. Furthermore, we acknowledge that the adopted instrumental-variables approach, despite constituting a significant methodological advance in the evaluation of IMF programmes, cannot guarantee unbiased causal estimates. However, our sensitivity analysis suggests that an unusual amount of unmeasured confounding would be required to cast serious doubt on our principal results. Finally, as already noted, we are not in a position to empirically substantiate the precise causal mechanisms at work, but we find that our aggregate findings are plausible in light of earlier scholarship, and we envisage that future investigations will be able to examine such mechanisms in greater detail.

To our knowledge, this is the first empirical study of the causal relation between structural adjustment and all-cause mortality rates or disability-adjusted life years. Our analysis withstands a range of sensitivity checks and adds to the scientific understanding of the distal causes of avoidable disease and death. It reveals that IMF-mandated policy reforms, especially those geared towards rapid privatisation, are harmful to population health and contribute to the persistence of high mortality and disease burdens in developing contexts. The role of international multilateral organisations such as the IMF in shaping population health outcomes should be taken seriously by researchers and policy-makers alike.

## Data Availability

Available from lead author upon request.

